# Pulmonary Vascular Disease in Veterans with Post-Deployment Respiratory Syndrome

**DOI:** 10.1101/2023.05.15.23289956

**Authors:** Sergey S. Gutor, Bradley W. Richmond, Vineet Agrawal, Evan L. Brittain, Matthew F. Mart, Ciara M. Shaver, Pingsheng Wu, Taryn K. Boyle, Ravinder R. Mallugari, Katrina Douglas, Robert N. Piana, Joyce E. Johnson, Robert F. Miller, John H. Newman, Timothy S. Blackwell, Vasiliy V. Polosukhin

## Abstract

**Background:** Increased frequency of exertional dyspnea has been documented in U.S. military personnel after deployment to Southwest Asia and Afghanistan. We studied whether continued exertional dyspnea in this patient population is associated with pulmonary vascular disease (PVD).

**Methods:** We recruited five Iraq and Afghanistan Veterans with post-deployment respiratory syndrome (PDRS) and continued exertional dyspnea to undergo a detailed clinical evaluation including symptom questionnaire, pulmonary function testing (PFT), surface echocardiography, and right heart catheterization (RHC) with exercise. We then performed detailed histomorphometry of blood vasculature in 52 Veterans with PDRS, 13 patients with advanced idiopathic pulmonary arterial hypertension (PAH) and 15 non-diseased (ND) control subjects.

**Results:** All five Veterans involved in clinical follow-up study had a continued dyspnea at exertion. On transthoracic echocardiography, we identified borderline or overt RV enlargement in three out of five Veterans. Right ventricle outflow tract (RVOT) acceleration time, a well-established surrogate measure of pulmonary pressure, was mildly reduced in three out of five Veterans. Of the five Veterans with PDRS who underwent RHC at exercise, we found that three had evidence of post-capillary PH at rest and one had PH at exercise. Morphometric evaluation of lung biopsy samples showed mild/moderate increase of fractional thicknesses of intima and media, and significant fibrosis of adventitia in pulmonary arteries in Veterans with PDRS compared to ND controls and PAH patients. Veterans with PDRS did not display plexiform or dilation/angiomatoid lesions, specific for PAH. Pulmonary veins showed similar levels of intima and adventitia fractional thickening in Veterans with PDRS and PAH patients compared to ND controls. In Veterans, IA veins were characterized by marked fibrous intima and adventitia thickening, usually with increased thickening and formation of multiple layers of elastic laminae, but without features of luminal occlusion, muscular hyperplasia or dilation/angiomatoid lesions seen in pulmonary veno-occlusive disease or chronic thromboembolic PH.

**Conclusions:** Our studies suggest that vasculopathy and PVD may explain exertional dyspnea and exercise limitation in some Veterans with PDRS. Evaluation for PVD should be considered in Iraq and Afghanistan Veterans with unexplained dyspnea.

## Introduction

Increased frequency of exertional dyspnea has been documented in U.S. military personnel after deployment to Southwest Asia and Afghanistan.^1–3^ Cohort studies have shown a wide spectrum of identifiable lung diseases in deployed Veterans (such as asthma and laryngeal disorders), yet up to 1/3 have been classified as undiagnosed exertional dyspnea.^4^ In 2021, we reported comprehensive analysis of open lung biopsy samples from 50 Veterans with exertional dyspnea unexplained by standard non-invasive testing and found complex pathological changes involving all distal lung compartments.^5^ We described chronic intrapulmonary lymphocytic inflammation, small airways pathology consistent with constrictive bronchiolitis (ConB), hypertensive-type pulmonary vasculopathy, diffuse interstitial fibrosis of alveolar tissue, and fibrosis involving visceral pleura. We proposed post-deployment respiratory syndrome (PDRS) as an umbrella term to define this complex of exposures, symptoms, and lung pathology.^5^

The dominance of exertional dyspnea and presence of hypertension-type vascular pathology, in the absence of substantial pulmonary function testing (PFT) abnormalities, in affected Veterans raise the possibility that pulmonary vascular disease (PVD) could account for exercise limitation in these patients. To determine whether Veterans with biopsy-proven PDRS and continued exertional dyspnea might have evidence of PVD, we recruited five Veterans (11-16 years after lung biopsy) to undergo a detailed clinical evaluation including symptom questionnaire, PFT, surface echocardiography, and right heart catheterization (RHC) with exercise. We also performed detailed histomorphometry of blood vasculature on lung biopsies from a larger group of subjects with PDRS to better define the pathological basis of PVD in this cohort.

## Materials and Methods

### Study design

We recruited five Iraq and Afghanistan Veterans to participate in a follow-up clinical study. All five Veterans were randomly chosen from previously described cohort^5, 6^ and were originally identified as having ConB^6^ and met criteria of PDRS^5^ based on clinical assessment and lung biopsy examination. In these Veterans, we performed standardized self-completed questionnaire for measuring impaired health, PFT, surface echocardiography, and RHC.

We performed comprehensive histopathological and morphometrical evaluation of intrapulmonary blood vessels in 52 Veterans with PDRS in which exertional dyspnea and exercise limitation were associated with deployment to Iraq and Afghanistan. We used lung biopsies from 50 individuals involved in our previous report.^5^ In addition, we included lung biopsies from two Veterans who participated in our clinical follow-up but were not evaluated in our prior publication. Lung biopsies were obtained by video-assisted thoracoscopic surgery (VATS) at Vanderbilt University Medical Center (VUMC) after ruling out identifiable cardiopulmonary disease by extensive non-invasive testing. All these individuals had pathological findings with no evidence of distinct lung pathologies, such as granulomatous diseases, interstitial lung disease, or chronic obstructive pulmonary disease, to account for their symptoms. Lung tissue specimens obtained from 15 non-diseased (ND) subjects and 13 end-stage patients with pulmonary arterial hypertension (PAH, Group 1 PH) were used as negative and positive controls, respectively. Specimens from patients with advanced PAH were obtained from explanted lungs at the time of lung transplantation. Lung tissue specimens from the ND group were obtained from the lungs rejected for lung transplantation as previously reported.^5^

### Demographics

As shown in **Table 1**, Veterans with PDRS were predominantly young (median age 36 at the time of biopsy), males, and non-smokers. All Veterans had exertional dyspnea and exercise intolerance as the main respiratory symptoms. All affected Veterans reported exposure to environmental hazards during deployment. Veterans reported inhalation exposures to burn pits, dust storms, diesel exhaust, human waste, combat smoke, and smoke from a sulfur mine fire.^5, 6^

**Table 1.** Demographic and clinical characteristics of study participants.

| Characteristics | ND Controls<br>(N=15) | Veterans with PDRS<br>(N=52) | Patients with PAH<br>(N=13) |
| --- | --- | --- | --- |
| **Age** (years) | 39 (21-50) | 36 (25-55) | 32 (15-56) |
| **Gender** |  |  |  |
| Male | 11 (73.3%) | 49 (94.2%) | 1 (7.8%) |
| Female | 4 (26.7%) | 3 (5.8%) | 12 (92.2%) |
| **Smoking history** |  |  |  |
| Lifelong non-smokers | 15 (100%) | 34 (65.4%) | 11 (84.6%) |
| Current smokers | — | 3 (5.8%) | — |
| Former smokers | — | 15 (28.8%) | 2 (15.4%) |
| **Reported exposures** |  |  |  |
| Sulfur dioxide | — | 24 (46.2%) | — |
| Burn pits | — | 22 (42.3%) | — |
| Dust storms | — | 9 (17.3%) | — |
| Burning oil, exhaust | — | 3 (5.8%) | — |
| Other (chemicals, industrial or combat smoke) | — | 8 (16%) | — |
| **Pulmonary function testing** |  |  |  |
| Normal | — | 31 (59.6%) | — |
| Low DLCO (<80% predicted) | — | 17 (32.7%) | — |
| Obstructive (FEV<sub>1</sub> <80% predicted, FEV<sub>1</sub>/FVC <0.7) | — | 3 (5.8%) | — |
| Restrictive (TLC <80% predicted) | — | 4 (7.7%) | — |
| **Computed tomography** |  |  |  |
| Normal | — | 28 (53.8%) | — |
| Mild/moderate air trapping | — | 20 (38.4%) | — |
| Basilar scarring | — | 2 (3.8%) | — |
| Mild interstitial fibrosis | — | 1 (1.9%) | — |
| Micronodules | — | 1 (1.9%) | — |
| Peripheral and ground-glass infiltrate | — | 2 (3.8%) | — |
| Emphysema | — | 2 (3.8%) | — |

### Institutional Review Board (IRB)

Histomorphometrical analysis was performed under the following Vanderbilt IRB approvals: 1) for surgical lung biopsy specimens from Veterans, #161046; 2) for patients with PAH who underwent lung transplantation, #060165; 3) lung tissue from deceased organ donors whose lungs were rejected for transplantation were exempt from IRB review. All clinical studies were approved by the Vanderbilt IRB (#200473 and #210973).

### Clinical follow-up evaluation

#### St. George Respiratory Questionnaire (SGRQ)

To quantify the degree of disability, recruited Veterans underwent the St. George’s Respiratory Questionnaire (SGRQ) which measures respiratory symptoms, activities limited by symptoms, and the impact of symptoms on well-being and social functioning.^7, 8^

#### Pulmonary Function Testing

PFT included spirometry, lung volumes, and diffusing capacity for carbon monoxide (DLCO) performed according to American Thoracic Society and European Respiratory guideline.^9^

#### Echocardiography

Echocardiograms were performed on a Philips iE33 machine (Netherlands) according to American Society of Echocardiography guidelines.^10, 11^ Data were analyzed by a board certified echocardiographer (Evan Brittain) using Tomtec software (Germany).

#### Exercise right heart catheterization and invasive cardiopulmonary exercise testing

Right heart catheterization was performed per clinical guideline as previously described.^12^ Briefly, in the supine position and under sterile conditions utilizing Seldinger technique, a 7-8Fr sheath was inserted under ultrasound guidance into the right internal jugular vein. A 7Fr Swan-Ganz pulmonary artery catheter (Edwards Life Sciences, Irvine, CA) was inserted through the sheath, balloon inflated, and sequentially advanced under fluoroscopic and hemodynamic guidance into the right atrium, right ventricle, pulmonary artery, and pulmonary wedge positions. Pressure measurements were zero balanced at the level of the right atrium while supine, and all pressure measurements at rest were obtained at end-expiration in mmHg. Cardiac output (CO) was estimated by the thermodilution method. Three Veterans underwent supine exercise RHC and two Veterans underwent invasive cardiopulmonary exercise testing (CPET), which consisted of seated upright RHC and gas exchange measurement. Research participants were subjected to a progressive load exercise protocol with a cycle ergometer either in the supine or seated upright position (Lode B.V., Groningen, Netherlands). For participants who underwent supine exercise, repeat hemodynamic measurements were made at various loads of exercise until the participant had reached symptom-limited peak exercise. Cardiac output was assessed by thermodilution at rest and peak exercise. For participants who underwent invasive CPET, a metabolic cart (MGC Diagnostics, St. Paul, MN, USA) was utilized to monitor gas exchange. Adequate test effort on invasive CPET was determined by respiratory exchange ratio (RER) via metabolic cart, and the exercise protocol continued until the participant was symptom limited. Hemodynamic measurements were made at various loads of exercise, and cardiac output was assessed by the Fick method utilizing measured VO2 from the metabolic cart. After exercise, participants were allowed to recover, and all catheters and sheaths were removed. Exercise RHC and invasive CPET were performed and interpreted by board-certified cardiologist (Vineet Agrawal and Robert Piana). CPET data were analyzed by Mathew Matt according to recently published recommendations.^13^

### Histopathological analysis and morphometry

Four serial paraffin tissue sections (5 μm) were cut from the same tissue blocks used in previous studies.^5^ Tissue sections were used for hematoxylin and eosin (H&E), periodic acid-Schiff (PAS), Verhoeff-Van Gieson (VVG) or PicroSirius Red stains. General manifestations of intrapulmonary inflammation and airway and alveolar tissue fibrosis were assessed on H&E, PAS and PicroSirius Red stained tissue sections, whereas vascular remodeling was assessed on VVG stained tissue sections. Morphometric analysis of blood vessels was performed at the three distinct branching levels including: 1) arteries from bronchovascular bundles with 300-500 μm in diameter (BVB arteries), 2) intra-acinar arteries with 70-150 μm in diameter (IA arteries), and 3) intra-acinar veins with 70-150 μm in diameter (IA veins). IA vessels, which are located within alveolar tissue and not associated with distal bronchioles, were considered as IA arteries or veins. Vessels with presence of internal elastic lamina were considered as IA arteries, whereas vessels with indistinct or absent internal elastic lamina were considered as IA veins.^14, 15^ Fractional thicknesses of intima, media and adventitia were measured in arteries, whereas fractional thicknesses of intima and adventitia were measured in veins. The areas of intima, media and adventitia were measured for all cross-sectioned BVB arteries and at least 10 randomly chosen IA arteries on digital images using ×10 or ×20 objectives depending on size. To determine intima fractional thickness, area of intima was normalized to the length of internal elastic lamina. To determine media and adventitia fractional thicknesses, areas of media or adventitia were normalized to the length of external elastic lamina. Data were presented as V:SA^Intima^, V:SA_Media_ or V:SA_Adventitia_, respectively. Fractional thicknesses of intima and adventitia were measured in at least 10 randomly chosen IA veins on digital images using ×20 objective. V:SA_Intima_ and V:SA_Adventitia_ in these veins were measured as intima or adventitia areas with following normalization to elastic lamina length. All morphometric measurements were done using a computerized image analyzer system (Image-Pro Plus, Media Cybernetics) by investigators (Sergey Gutor and Vasiliy Polosukhin) who were blinded to the study groups.

### Statistical analysis

Demographic, clinical, morphological, and physiological data are reported as median (range) or mean ± standard deviation (SD) according to their distributions. For categorical variables, proportions were used. Comparisons between symptomatic Veterans, PAH patients and ND controls were conducted using t-test or Mann-Whitney U-test for continuous variables. Follow-up data were compared with initial values by paired t-test or paired Wilcoxon test according to distribution. Hierarchical cluster analysis was used to identify and differentiate subgroups based on pathological patterns.^16^ A hierarchical cluster tree (dendrogram) was constructed based on Euclidean distance. Subgroups were formed in an agglomerative manner, starting with each participant as his/her own subgroup and pairing the two closest subgroups together at every step until only one group of all participants remained. We applied the Gap statistic and Total Within Sum of Square statistic method for determination of the optimal number of clusters.^17^ The number of subgroups was also confirmed by a visual inspection of the dendrogram and a Multi-Dimensional Scaling plot. All analyses were performed using R-software version 3.5.2 (www.r-project.org).

## RESULTS

### Clinical follow-up in Veterans with PDRS

We recruited 5 Veterans who had previously undergone lung biopsy for unexplained exertional dyspnea (11-16 years prior to follow-up) and had findings consistent with PDRS after an extensive clinical evaluation and lung biopsy examination. Veteran participants were all males, ages 41-62, and non-smokers. All had a continued dyspnea at exertion and high symptom burden on the St. George Respiratory Questionnaire (SGRQ), which measures respiratory symptoms, activities limited by symptoms, and the impact of symptoms on well-being and social functioning. Four Veterans had normal PFT parameters, and one had mild obstruction. Of note, this Veteran had mild obstruction at his initial assessment (at the time of biopsy) without further decline in PFT parameters (**Table 2**).

**Table 2.** Clinical data for Veterans undergoing right heart catheterization (N=5).

|  | Case 1 | Case 2 | Case 3 | Case 4 | Case 5 | Interpretation |
| --- | --- | --- | --- | --- | --- | --- |
| Time period between biopsy and current assessment (years) | 16 | 16 | 11 | 11 | 14 |  |
| **Symptoms** |  |  |  |  |  |  |
| SGRQ |  |  |  |  |  |  |
| Symptoms | 93 | 87 | 48 | 83 | 56 | The SGRQ scores ranges from 0 to 100 where 100 indicates the worst quality of life<sup>7,8</sup> |
| Activity | 80 | 87 | 57 | 100 | 45 |  |
| Impacts | 88 | 53 | 18 | 77 | 32 |  |
| Total | 87 | 69 | 34 | 85 | 41 |  |
| **Pulmonary Function Tests** |  |  |  |  |  |  |
| FEV<sub>1</sub>/FVC | 0.67 | 0.78 | 0.78 | 0.73 | 0.78 | < 0.7 indicates airflow obstruction<sup>9</sup> |
| FEV<sub>1</sub> (% predicted) | 88 | 94 | 112 | 82 | 100 | Indicates the severity of airflow obstruction<sup>9</sup> |
| FVC (% predicted) | 103 | 93 | 109 | 85 | 91 |  |
| TLC (% predicted) | 111 | 105 | 108 | 97 | 96 | <80 indicates airflow restriction<sup>9</sup> |
| DLCO (% predicted) | 111 | 104 | 105 | 103 | 152 | <60 indicates decreased diffusing lung capacity<sup>9</sup> |
| **Echocardiography** |  |  |  |  |  |  |
| **Left heart** |  |  |  |  |  |  |
| Left ventricular ejection fraction (%) | 64.7 | 57.6 | 62 | 62.6 | 67.4 | 50% to 70% indicates norm<sup>10,11</sup> |
| Left atrial volume index (mL/m<sup>2</sup>) | 30.5 | – | 23.8 | 20.4 | 13.5 | <34 indicates norm<sup>10,11</sup> |
| E/e' peak/mean | 8.9/7.5 | 5.2/5.4 | 8.4/7.5 | 15/13.5 | 9.9/8.4 | >15 indicates increased filling pressure<sup>10,11</sup> |
| Tricuspid regurgitation velocity (m/sec) | 1.29 | 1.81 | – | – | 2.16 | >2.8 indicates diastolic dysfunction<sup>10,11</sup> |
| **Right heart** |  |  |  |  |  |  |
| RV Basal Dimension A4C (mm) | 43 | 42 | 31 | 45 | 39 | >42 indicates RV dilatation<sup>21-23</sup> |
| TAPSE (cm) | 2.0 | 2.3 | 2.3 | 2.4 | 2.3 | <1.7 indicates RV systolic dysfunction<sup>20</sup> |
| Tricuspid Annular S' (cm/s) | 17 | 17 | 18 | 17 | 17 | <10 indicates RV systolic dysfunction<sup>21-23</sup> |
| Fractional Area Change (%) | 40.1 | 43.1 | 39.7 | 44.4 | 48.6 | <35 indicates RV systolic dysfunction<sup>21-23</sup> |
| RVOT acceleration time (ms) | 138 | 144 | 114 | 128 | 96 | <100 - highly suggestive of PH, 100-130 – borderline<sup>21-23</sup> |
| **RHC Parameters** |  |  |  |  |  |  |
| **Resting (supine)** |  |  |  |  |  |  |
| mPAP (mmHg) | 21 | 26 | 16 | 23 | 18 | >20 indicates PH<sup>19</sup> |
| PAWP (mmHg) | 8 | 18 | 13 | 14 | 16 | > 15 indicates left sided congestion<sup>19</sup> |
| CO (L/min) | 8.4 | 5.4 | 6.6 | 5.5 | 4.9 |  |
| PVR (mmHg·min/L) | 1.55 | 1.48 | 0.45 | 1.64 | 0.41 |  |
| **Exercise RHC / Invasive CPET\*** |  |  |  |  |  |  |
| Peak workload (Watts) | 120 | 213 | 80 | 60 | 225 |  |
| Peak VO<sub>2</sub> (mL/kg/min) | – | 28.7 | – | – | 20.8 |  |
| VO<sub>2</sub> max predicted (%) | – | 93 | – | – | 67 |  |
| Anaerobic threshold (mL/kg/min) | – | 21.8 | – | – | 14.3 |  |
| Anaerobic threshold predicted (%) | – | 71 | – | – | 37 | <40 indicates reduced anaerobic threshold<sup>18</sup> |
| mPAP (mmHg) | 31 | 35 | 44 | 40 | 32 |  |
| PAWP (mmHg) | 26 | 22 | 24 | 26 | 28 | >25 indicates exercise HFpEF<sup>19</sup> |
| CO (L/min) | 16.4 | 17.9 | 12.8 | 12.4 | 23.8 |  |
| mPAP/CO slope (mmHG/L/min) | 1.25 | 1.55 | 4.52 | 2.46 | 0.74 | >3 indicates exercise PH<sup>19</sup> |
| PAWP/CO slope (mmHG/L/min) | 2.25 | 1.63 | 1.77 | 1.74 | 0.63 | >2 indicates exercise HFpEF<sup>19</sup> |

\* – Exercise RHC was performed in three Veterans (Cases 1,3 and 4); Invasive CPET was performed in two Veterans (Cases 2 and 5).
NS – lifelong non-smokers; FS – former smokers; RV – right ventricle; TAPSE – tricuspid annular plane systolic excursion; Tricuspid Annular S' – tricuspid annular peak systolic velocity; RVOT – RV outflow tract; mPAP – mean pulmonary arterial pressure; CO – cardiac output; PCWP – pulmonary capillary wedge pressure; PVR – pulmonary vascular resistance; CPET – cardio-pulmonary exercise test; mPAP/CO slope calculated by formula $(mPAP_{\text{exercise}} - mPAP_{\text{rest}}) / (CO_{\text{exercise}} - CO_{\text{rest}})$ ; PAWP/CO slope calculated by formula $(PAWP_{\text{exercise}} - PAWP_{\text{rest}}) / (CO_{\text{exercise}} - CO_{\text{rest}})$ ; pc-PH – post-capillary pulmonary hypertension; HFpEF – heart failure with preserved ejection fraction. Red indicates values outside the normal range.

Veterans underwent echocardiography and exercise RHC or invasive CPET, which included exercise RHC with a metabolic cart to determine anaerobic threshold. Four of the five Veterans achieved submaximal workloads (cases 1,3,4, and 5), while one Veteran (Case 2) achieved 105% predicted workload. Of the two patients who underwent invasive CPET, one Veteran (Case 5) had reduced peak VO_2_ (67% predicted) with a reduced anaerobic threshold (37% of predicted VO_2_ max)^18^ consistent with a primary cardiocirculatory limitation and one Veteran (Case 2) had a normal peak VO_2_ (93% predicted) and normal anaerobic threshold.

We defined resting and exercise PH according to the 2022 European Society of Cardiology/European Respiratory Society (ESC/ERS) Guidelines for the Diagnosis and Treatment of PH.^19^ Resting PH, defined as a mean pulmonary arterial pressure (mPAP) > 20 mmHg, was present in three out of five Veterans (Cases 1, 2, and 4). Two Veterans (Cases 2 and 5) showed evidence of mildly increased pulmonary artery wedge pressures at rest (PAWP) > 15 mmHg, suggestive of left heart congestion. With peak exercise, one Veteran (Case 3) demonstrated exercise PH with mPAP/CO slope > 3 mmHg/L/min and three Veterans demonstrated evidence of moderate isolated post-capillary PH (Cases 1, 4, 5) and met criteria for heart failure with preserved ejection fraction (HFpEF), which is defined by either an exercise PCWP > 25 mmHg or PCWP/CO slope of > 2 mmHg/L/min (**Table 2**).

On transthoracic echocardiography, we identified borderline or overt RV enlargement in three Veterans (60%).^20^ RVOT acceleration time, a well-established surrogate measure of pulmonary pressure,^21–23^ was mildly reduced in three out of five Veterans (Cases 3-5). Echocardiographic measures of left ventricular size and systolic and diastolic function were normal (**Table 2**). Together, these findings suggest that some Veterans with PDRS in our study have mild/moderate PH that is predominately post-capillary, and/or HFpEF, which could explain their continuing exertional dyspnea.

### Pathological evaluation

Microscopic re-evaluation of lung biopsy sections from Veterans with PDRS confirmed our previous findings of intrapulmonary inflammation and multicompartmental lung pathology.^5, 24^ In this study, we focused on vascular pathology. In BVB arteries, pathological changes were characterized by mild-to-moderate media thickening due to smooth muscle hyperplasia/hypertrophy, and fibrosis of adventitia, usually without thickening. Neointimal formation was variable and often had eccentric, cushion-like localized manifestations (**Figure 1A**). Pathological changes were not limited by BVB arteries but extended to distal vessel generations located within alveolar tissue. IA arteries were characterized by moderately increased intima and media thicknesses and significant fibrosis and thickening of adventitia. Neointimal lesions were accompanied by moderate-to-severe fibrosis (**Figure 2A**). Some arteries (∼ 5-10%) showed laminar fibrosis of the intima due to the concentrically arranged fibrous layers leading to partial obstruction (also known as “onion-skin lesions”). In comparison to PAH patients, Veterans with PDRS had more prominent fibrosis at all generations of pulmonary arteries analyzed (**Figures 1, 2**). Comparison analysis of morphometric parameters showed a mild increase of fractional thickness of intima and moderate increase of media in both BVB and IA arteries in Veterans with PDRS compared to ND controls and PAH patients. Fractional thickening of adventitia was similar in BVB arteries in all study groups, but gradually increased in IA arteries in Veterans with PDRS and PAH patients compared to ND controls (**Figures 1 and 2**). In contrast to PAH patients, Veterans with PDRS did not display plexiform or dilation/angiomatoid lesions.

**Figure 1.**
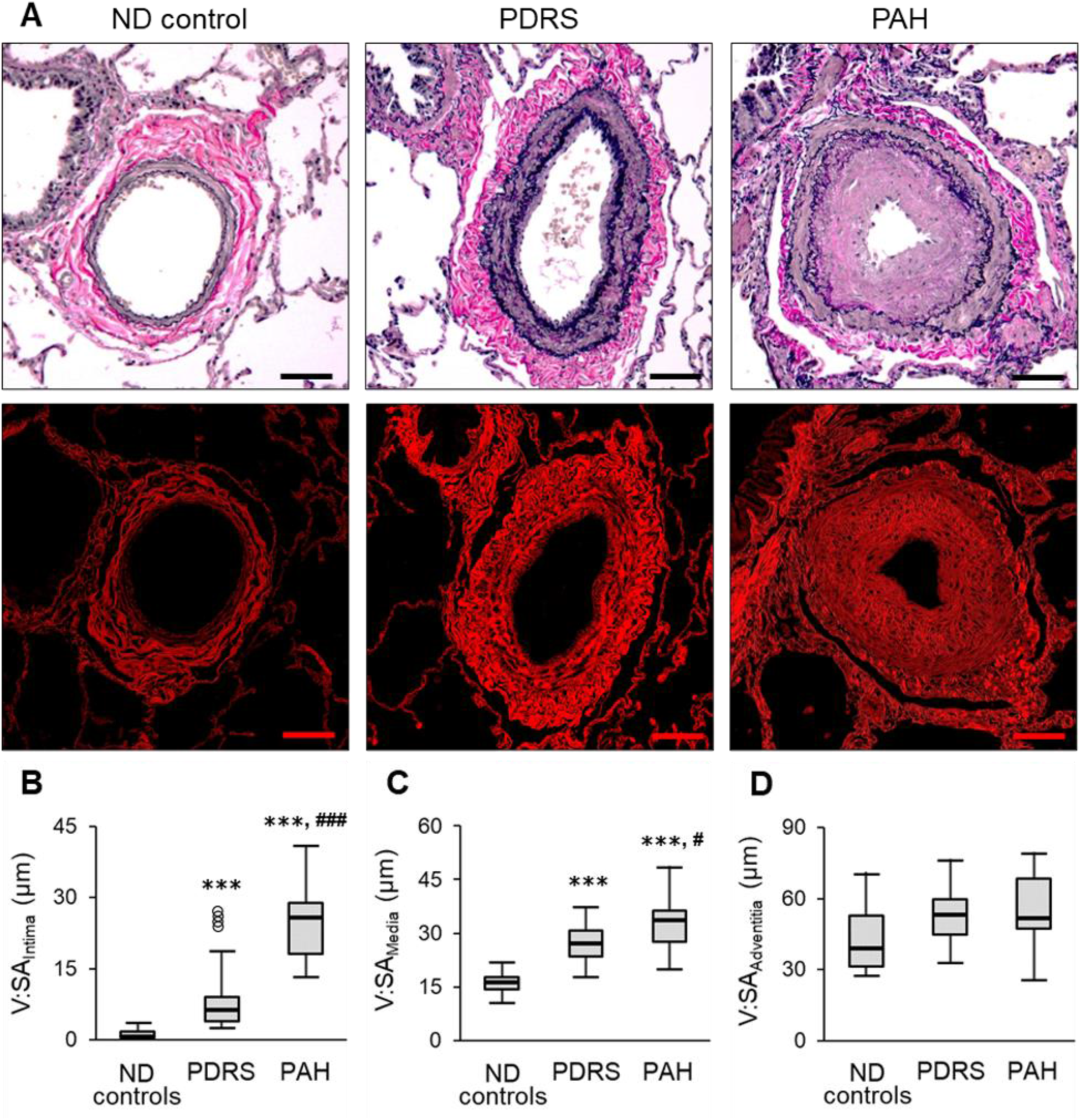
Histopathology of BVB arteries. Representative images of BVB arteries. A – Top row – normal-appearing artery from ND subject; mild neointimal formation (black arrow) and moderate muscularization in artery from Veteran with PDRS; prominent intima and moderate media thicknesses in artery from PAH patient. Verhoeff-Van Gieson (VVG) staining. Bottom row – concentric organization of fine collagen fibers in adventitia in artery from ND control; marked collagen deposits in media and adventitia in artery from Veteran with PDRS; moderate media fibrosis in artery from PAH patient. PicroSirius red staining; collagen red fluorescence. Scale bars = 100 µm. B-D – Graphs showing progressively increased fractional thicknesses of intima (B), media (C) or adventitia (D) in PDRS and PAH compared to ND controls. Data presented as V:SA_Intima_ for intima thickness, V:SA_Media_ for media thickness or V:SA_Adventitia_ for adventitia thickness. Boxes represent the interquartile range, whiskers extend to the most extreme data point which is no more than 1.5 times the interquartile range from the box, and circles beyond the whiskers are extreme values, the line within the box represents the median. Groups were compared pairwise using Mann-Whitney U or Student t tests depending on variable distribution. Estimated p-values were Bonferroni-adjusted. **\*\*\*** - p<0.001 compared to ND controls; **#** - p<0.05 compared to PDRS; **###** - p<0.001 compared to PDRS.

**Figure 2.**
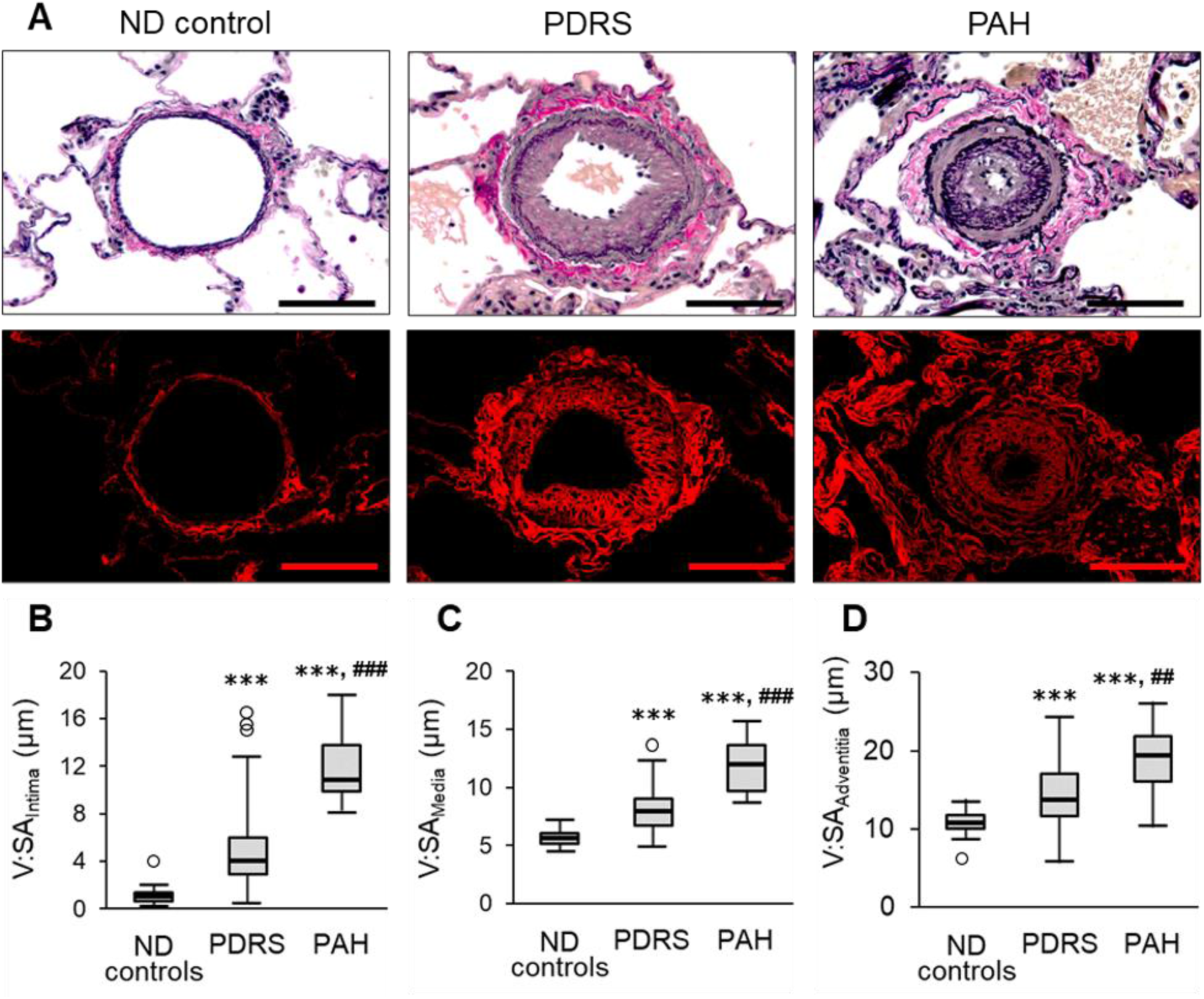
Histopathology of IA arteries. Representative images of IA arteries. A – Top row – normal-appearing artery from ND subject; moderate neointimal formation and moderate muscularization in artery from Veteran with PDRS; “onion-skin” laminar neointimal lesion with almost complete luminal narrowing and moderate media thicknesses in artery from PAH patient. Verhoeff-Van Gieson (VVG) staining. Bottom row – fine collagen fibers organization in artery from ND control; marked collagen deposits in intima and adventitia in artery from Veteran with PDRS; mild intima fibrosis in artery from PAH patient. PicroSirius red staining; collagen red fluorescence. Scale bars = 50 µm. B-D – Graphs showing progressively increased fractional thicknesses of intima (B), media (C) or adventitia (D) in PDRS and PAH compared to ND controls. Data presented as V:SA_Intima_ for intima thickness, V:SA_Media_ for media thickness or V:SA_Adventitia_ for adventitia thickness. Boxes represent the interquartile range, whiskers extend to the most extreme data point which is no more than 1.5 times the interquartile range from the box, and circles beyond the whiskers are extreme values, the line within the box represents the median. Groups were compared pairwise using Mann-Whitney U or Student t tests depending on variable distribution. Estimated p-values were Bonferroni-adjusted. **\*\*\*** – p<0.001 compared to ND controls; **##** – p<0.01 compared to PDRS; **###** – p<0.001 compared to PDRS.

Although end-stage PAH patients showed more severe remodeling in BVB and IA arteries, IA veins showed similar levels of intima and adventitia fractional thickening in Veterans with PDRS and PAH patients compared to ND controls (**Figure 3A-C**). However, in Veterans, IA veins were characterized by marked fibrosis of intima and adventitia, usually with increased thickening and formation of multiple layers of elastic laminae, but without features of luminal occlusion, muscular hyperplasia or dilation/angiomatoid lesions seen in pulmonary veno-occlusive disease or chronic thromboembolic PH.^25–27^

**Figure 3.**
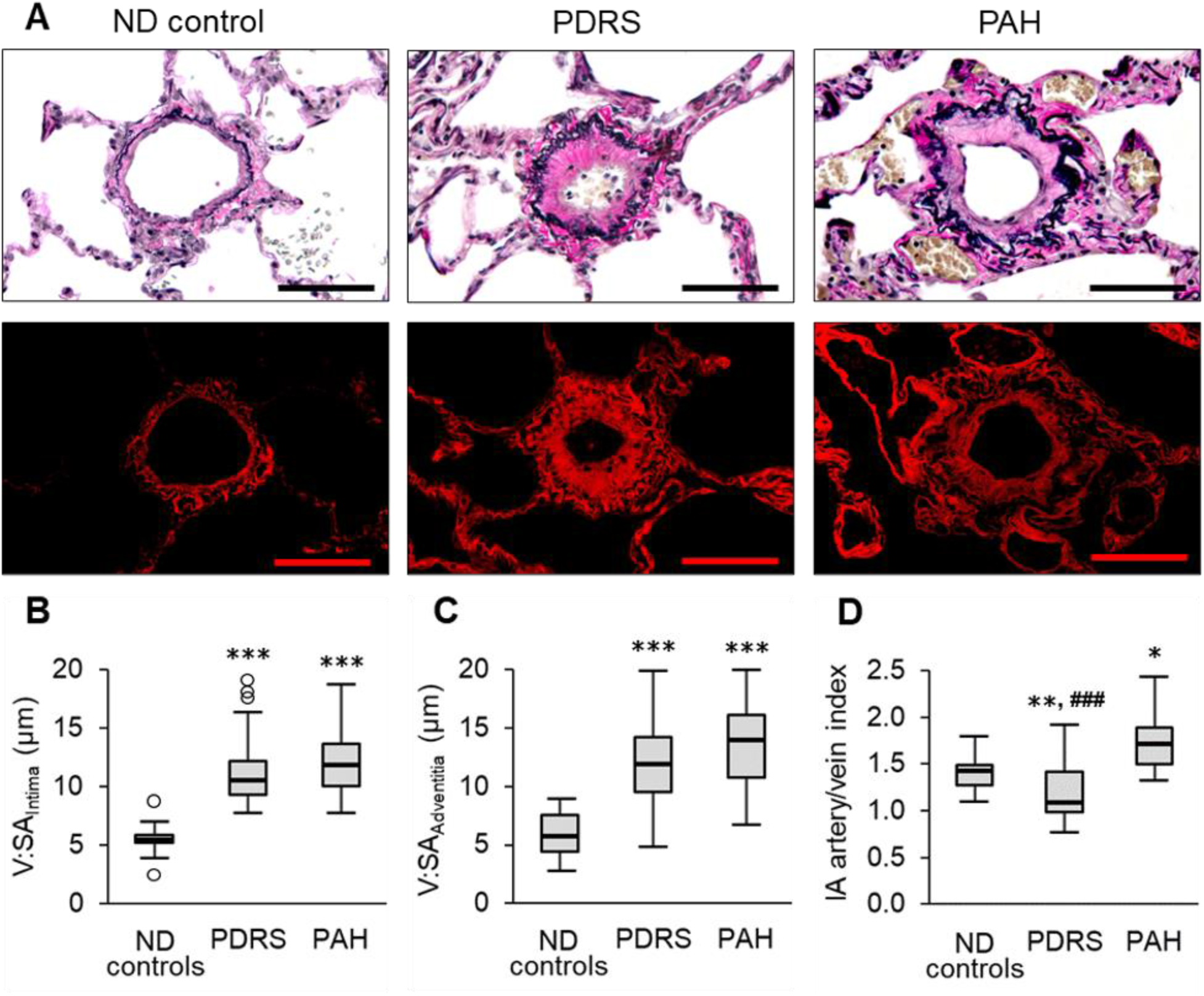
Histopathology of IA veins. Representative images of IA veins. A – Top row – normal-appearing vein from ND subject; intima and adventitia thickening in vein from PDRS and PAH compared to ND control. Verhoeff-Van Gieson (VVG) staining. Bottom row – marked collagen depositions in intima and adventitia in vein from Veteran with PDRS compared to both ND controls and PAH patients. PicroSirius red staining; collagen red fluorescence. Scale bars = 50 µm. B, C – Graphs showing progressively increased fractional thicknesses of intima (B) or adventitia C) in PDRS and PAH compared to ND controls. Data is presented as V:SA_Intima_ for intima thickness or V:SA_Adventitia_ for adventitia thickness. D – Graph showing modestly increased artery/vein wall thickness ratio in PAH patients compared to ND controls and significantly decreased artery/vein wall thickness ratio in PDRS compared to both ND controls and PAH patients. Data is presented as IA artery/vein index. Boxes represent the interquartile range, whiskers extend to the most extreme data point which is no more than 1.5 times the interquartile range from the box, and circles beyond the whiskers are extreme values, the line within the box represents the median. Groups were compared pairwise using Mann-Whitney U or Student t tests depending on variable distribution. Estimated p-values were Bonferroni-adjusted. **\*** – p< 0.05 compared to ND controls; **\*\*** – p< 0.01 compared to ND controls; **\*\*\*** – p<0.001 compared to ND controls; **##** - p<0.001 compared to PAH.

To determine which anatomical compartment, arterial or venular, is predominately involved in vessel pathology in PDRS, we calculated the ratio of wall thicknesses between IA pulmonary arteries and veins. This index was measured for each study participant as the mean of sum of intima, media and adventitia fractional thicknesses for arteries divided by the mean of sum of fractional thicknesses of intima and adventitia for veins. **Figure 3D** shows a modest increase in the artery/vein index in PAH patients compared to ND controls, while lungs from PDRS patients had a reduced artery/vein index compared to both ND controls and PAH patients. This finding indicates more prominent remodeling of pulmonary veins relative to arteries in PDRS, which is distinct from the predominant pre-capillary vessel pathology in PAH.

To further investigate whether pathological changes in the lung vasculature in Veterans with PDRS were distinct from ND controls and PAH patients, we performed unsupervised hierarchical clustering using all morphometric parameters. This approach showed that the optimal number of clusters was 3 (**Supplemental Figure 1**) and separated ND controls and PAH patients; however, some Veterans were incorporated into the clusters containing ND controls or PAH patients. The dendrogram in **Figure 4** shows that the 1^st^ cluster combines all ND controls and 16 Veterans; the 2^nd^ cluster consists of Veterans only; and the 3^rd^ cluster combines 10 Veterans and all PAH patients. To confirm whether this distribution reflects mild, moderate, and severe vascular pathology, we grouped all morphometric parameters from Veterans according to clusters and performed comparison analysis between clusters. We found a significant increase of all measured parameters across the three patient clusters, consistent with increased pathological remodeling in all vessel wall components (**Supplemental Figure 2**). Secondary analysis of morphometric parameters from the five Veterans who participated in the clinical follow-up study showed that pathological remodeling in three (Cases 2, 4 and 5) fell into the least affected cluster (cluster 1), while two Veterans (Cases 1 and 3) were grouped in the moderately affected cluster (cluster 2).

**Figure 4.**
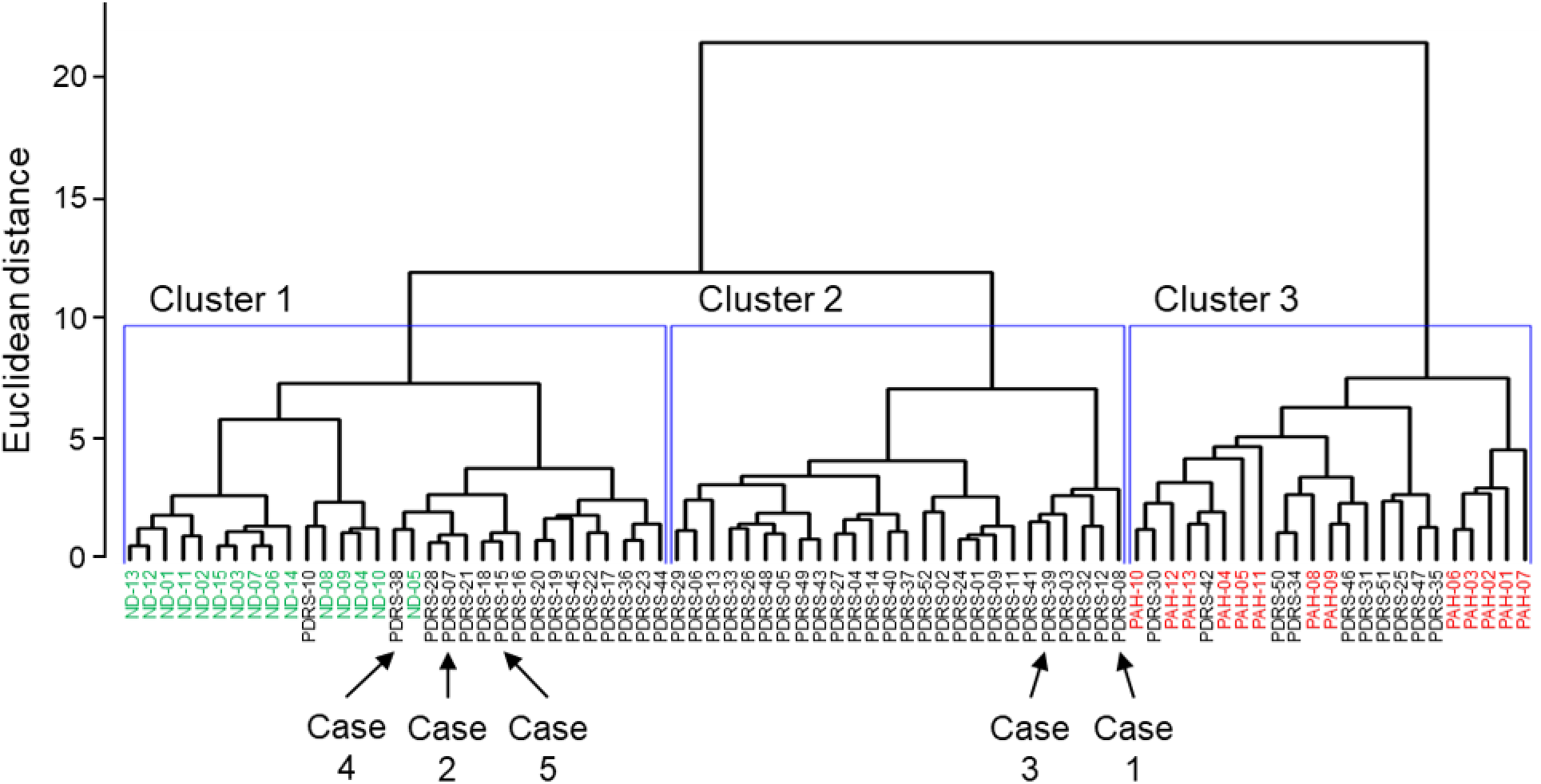
Unsupervised hierarchical clustering analysis. Unsupervised Hierarchical Clustering Dendrogram using Euclidean distance metric for clustering and calculating the similarity between two clusters with Ward’s method.^18^ Arrows for Cases 1-5 identify Veterans recruited for clinical follow-up study.

## DISCUSSION

This clinical follow-up study showed that all five Veterans with biopsy-proven PDRS had continued dyspnea at exertion and evidence of PVD. RHC showed that three of five Veterans had mild resting PH and a fourth had exercise PH. In all cases, PVR was < 2 Wood units, suggesting post-capillary PH. Although post-capillary PH is well-described in the setting of left heart failure,^28–30^ no participants in our study had echocardiographic evidence of heart failure with reduced or preserved ejection fraction (HFrEF or HFpEF). However, we observed marked fibrotic remodeling of intra-acinar veins in Veterans, which was relatively more severe than in PAH patients. Thus, we speculate that fibrous remodeling of pulmonary veins and resulting narrowing of the cross-sectional luminal area could contribute to the post-capillary PH observed in these Veterans. Although we found no echocardiographic evidence for diastolic dysfunction in our study, left-heart catheterization with assessment of the diastolic pressure-volume relationship will be needed to conclusively exclude diastolic dysfunction as a contributor to post-capillary PH in these Veterans.

We previously reported multicompartmental lung pathology with hypertensive-type changes in BVB arteries and capillary rarefaction in alveolar tissue were important pathological features of PDRS.^5^ In this study, we extend these findings to show that vascular pathology involves multiple vascular compartments (arteries, capillaries, and veins) and multiple anatomic levels (BVB arteries, IA arteries and veins). Generally, these findings are similar to, although less severe than, patients with idiopathic PAH who underwent lung transplantation. Together, our studies suggest that vasculopathy and subsequent PH may explain exertional dyspnea and exercise limitation in some Veterans who have an otherwise unremarkable non-invasive evaluation.

We speculate that PH in this cohort of Veterans with PDRS is associated with mixed inhalation exposures received during deployment.^5, 6^ Recent epidemiologic and animal studies have been demonstrated a causative relationship between inhalation of noxious substances and development of PH. Thus, PH has been reported in association with chronic exposure to tobacco and woodsmoke, air pollution, particulate matter with a diameter < 2.5 µm (PM_2.5_), and exposure to other chemicals, dust, gases or fumes exposures.^31–36^ Experimental works demonstrated that exposure to diesel exhaust or urban ambient PM_2.5_ is sufficient to induce marked pulmonary arterial thickening and increased right ventricle systolic pressure in mice, supporting a causal relationship between inhalation of noxious particles and the development of PH.^37–39^

Conceptually, vasculopathy in the setting of inhalation of noxious substances could result from “inside-out” or “outside-in” processes. In the inside-out model, absorption of toxins such as cigarette smoke or pollution into the bloodstream at the capillary/alveolar interface could result in direct damage to endothelial cells through well-established pathways involving oxidative stress and induction of endothelial cell dysfunction.^40–44^ Alternatively, vasculopathy could result from an “outside-in” process whereby inflammation from adjacent airways induced by inhalation of noxious particles causes pathologic remodeling of blood vessels. In support of this concept, the extent of perivascular inflammation in PAH has been shown to correlate with pulmonary hemodynamics, vascular remodeling, and clinical outcome.^45–48^ Persistent airway and parenchymal lymphocytic inflammation in association with pure Th1 adaptive immune response was defined as a central feature of PDRS that raises the possibility that the PVD could progress in this patient population over time.^5, 24^ The relative contributions of inside-out and outside-in pathology in each vascular compartment and anatomic level await further testing in relevant animal models.

Our study has several important limitations. First, because vasculopathy in these Veterans occurs in association with pathology in other lung compartments including small airways, the lung parenchyma, and the pleura, we cannot definitely link PVD with symptoms in individual Veterans. Emerging non-invasive techniques capable of interrogating multiple lung compartments simultaneously (such as hyperpolarized xenon MRI^49^) may help to assign the relative importance of pathology in various lung compartments in individual patients. Second, pathologic analyses were performed on archived biopsies obtained more than a decade prior to exercise RHC. Thus, we cannot ascertain whether PH was present at the time of biopsy or developed or progressed over time. Third, we performed exercise RHC in three Veterans using a multistage exercise protocol prior to developing the capability to perform invasive CPET using a continuous ramp exercise protocol. In these Veterans, exercise could not be standardized against anaerobic threshold, and thus the exercise level likely varied between these participants and those who performed invasive CPET due to differences in exercise protocol. Finally, we acknowledge that our case series of five patients is insufficient to conclude that development of PH in these patients is widespread. However, it is important to note that several of the Veterans who underwent RHC had relatively minor vasculopathy present on surgical lung biopsies compared to other PDRS patients, suggesting more prominent findings may exist in other Veterans with PDRS. Larger studies involving a group of Veterans with heterogenous civilian and deployment exposure profiles will be necessary to estimate the prevalence of PH in this population.

In summary, here we report complex vascular pathology in surgical lung biopsies from Iraq and Afghanistan Veterans with unexplained dyspnea accompanied by PVD in a subset. Our findings suggest PVD should be considered in the evaluation of Iraq and Afghanistan Veterans with unexplained dyspnea.

Median and range (minimum and maximum) are indicated for age. Number (percent) is indicated for gender and smoking history.

## Data Availability

All data produced in the present study are available upon reasonable request to the authors

## REFERENCES

1. Helmer DA, Rossignol M, Blatt M, Agarwal R, Teichman R, Lange G. Health and exposure concerns of veterans deployed to Iraq and Afghanistan. J Occup Environ Med. 2007;49:475–480. doi: 10.1097/JOM.0b013e318042d682

2. McAndrew LM, Teichman RF, Osinubi OY, Jasien JV, Quigley KS. Environmental exposure and health of Operation Enduring Freedom/Operation Iraqi Freedom veterans. J Occup Environ Med. 2012;54:665–669. doi: 10.1097/JOM.0b013e318255ba1b

3. Garshick E, Abraham JH, Baird CP, Ciminera P, Downey GP, Falvo MJ, Hart JE, Jackson DA, Jerrett M, Kuschner W, et al. Respiratory Health after Military Service in Southwest Asia and Afghanistan. An Official American Thoracic Society Workshop Report. Ann Am Thorac Soc. 2019;16:e1–e16. doi: 10.1513/AnnalsATS.201904-344WS

4. Morris MJ, Walter RJ, McCann ET, Sherner JH, Murillo CG, Barber BS, Hunninghake JC, Holley AB. Clinical Evaluation of Deployed Military Personnel With Chronic Respiratory Symptoms: Study of Active Duty Military for Pulmonary Disease Related to Environmental Deployment Exposures (STAMPEDE) III. Chest. 2020;157:1559–1567. doi: 10.1016/j.chest.2020.01.024

5. Gutor SS, Richmond BW, Du RH, Wu P, Sandler KL, MacKinnon G, Brittain EL, Lee JW, Ware LB, Loyd JE, et al. Postdeployment Respiratory Syndrome in Soldiers With Chronic Exertional Dyspnea. Am J Surg Pathol. 2021;45:1587–1596. doi: 10.1097/PAS.0000000000001757

6. King MS, Eisenberg R, Newman JH, Tolle JJ, Harrell FE, Jr., Nian H, Ninan M, Lambright ES, Sheller JR, Johnson JE, et al. Constrictive bronchiolitis in soldiers returning from Iraq and Afghanistan. N Engl J Med. 2011;365:222–230. doi: 10.1056/NEJMoa1101388

7. Jones PW, Quirk FH, Baveystock CM. The St George’s Respiratory Questionnaire. Respir Med. 1991;85 Suppl B:25-31; discussion 33-27. doi: 10.1016/s0954-6111(06)80166-6

8. Craig CL, Marshall AL, Sjostrom M, Bauman AE, Booth ML, Ainsworth BE, Pratt M, Ekelund U, Yngve A, Sallis JF, et al. International physical activity questionnaire: 12-country reliability and validity. Med Sci Sports Exerc. 2003;35:1381–1395. doi: 10.1249/01.MSS.0000078924.61453.FB

9. Pellegrino R, Viegi G, Brusasco V, Crapo RO, Burgos F, Casaburi R, Coates A, van der Grinten CP, Gustafsson P, Hankinson J, et al. Interpretative strategies for lung function tests. Eur Respir J. 2005;26:948–968. doi: 10.1183/09031936.05.00035205

10. Rudski LG, Lai WW, Afilalo J, Hua L, Handschumacher MD, Chandrasekaran K, Solomon SD, Louie EK, Schiller NB. Guidelines for the echocardiographic assessment of the right heart in adults: a report from the American Society of Echocardiography endorsed by the European Association of Echocardiography, a registered branch of the European Society of Cardiology, and the Canadian Society of Echocardiography. J Am Soc Echocardiogr. 2010;23:685–713. doi: S0894-7317(10)00434-7

11. Lang RM, Badano LP, Mor-Avi V, Afilalo J, Armstrong A, Ernande L, Flachskampf FA, Foster E, Goldstein SA, Kuznetsova T, et al. Recommendations for cardiac chamber quantification by echocardiography in adults: an update from the American Society of Echocardiography and the European Association of Cardiovascular Imaging. J Am Soc Echocardiogr. 2015;28:1–39. doi: S0894-7317(14)00745-7

12. Agrawal V, D’Alto M, Naeije R, Romeo E, Xu M, Assad TR, Robbins IM, Newman JH, Pugh ME, Hemnes AR, et al. Echocardiographic Detection of Occult Diastolic Dysfunction in Pulmonary Hypertension After Fluid Challenge. J Am Heart Assoc. 2019;8:e012504. doi: 10.1161/JAHA.119.012504

13. Glaab T, Taube C. Practical guide to cardiopulmonary exercise testing in adults. Respir Res. 2022;23:9. doi: 10.1186/s12931-021-01895-6

14. Townsley MI. Structure and composition of pulmonary arteries, capillaries, and veins. Compr Physiol. 2012;2:675–709. doi: 10.1002/cphy.c100081

15. Humbert M, Guignabert C, Bonnet S, Dorfmuller P, Klinger JR, Nicolls MR, Olschewski AJ, Pullamsetti SS, Schermuly RT, Stenmark KR, et al. Pathology and pathobiology of pulmonary hypertension: state of the art and research perspectives. Eur Respir J. 2019;53. doi: 10.1183/13993003.01887-2018

16. Murtagh F, Legendre P. Ward’s Hierarchical Agglomerative Clustering Method: Which Algorithms Implement Ward’s Criterion? J Classif. 2014;31:274–295. doi: 10.1164/rccm.201712-2513OC

17. Tibshirani R, Walther G, Hastie T. Estimating the number of clusters in a data set viathe gap statistic. J R Statist Soc B. 2001;63:411-423. doi: 10.1164/rccm.201712-2513OC

18. Balady GJ, Arena R, Sietsema K, Myers J, Coke L, Fletcher GF, Forman D, Franklin B, Guazzi M, Gulati M, et al. Clinician’s Guide to cardiopulmonary exercise testing in adults: a scientific statement from the American Heart Association. Circulation. 2010;122:191–225. doi: 10.1161/CIR.0b013e3181e52e69

19. Humbert M, Kovacs G, Hoeper MM, Badagliacca R, Berger RMF, Brida M, Carlsen J, Coats AJS, Escribano-Subias P, Ferrari P, et al. 2022 ESC/ERS Guidelines for the diagnosis and treatment of pulmonary hypertension. Eur Respir J. 2022. doi: 10.1183/13993003.00879-2022

20. Saxena N, Rajagopalan N, Edelman K, Lopez-Candales A. Tricuspid annular systolic velocity: a useful measurement in determining right ventricular systolic function regardless of pulmonary artery pressures. Echocardiography. 2006;23:750–755. doi: 10.1111/j.1540-8175.2006.00305.x

21. Parasuraman S, Walker S, Loudon BL, Gollop ND, Wilson AM, Lowery C, Frenneaux MP. Assessment of pulmonary artery pressure by echocardiography-A comprehensive review. Int J Cardiol Heart Vasc. 2016;12:45–51. doi: 10.1016/j.ijcha.2016.05.011

22. Aashish A, Giridharan S, Karthikeyan S, Ganesh BA, Prasath PA. Assessment of Pulmonary Artery Pressures by Various Doppler Echocardiographic Parameters and its Correlation with Cardiac Catheterization in Patients with Pulmonary Hypertension. Heart Views. 2020;21:263–268. doi: 10.4103/HEARTVIEWS.HEARTVIEWS_133_20

23. Gursel G, Ozdemir U, Guney T, Karaarslan N, Tekin O, Ozturk B. The usefulness of subxiphoid view in the evaluation of acceleration time and pulmonary hypertension in ICU patients. Echocardiography. 2020;37:1345–1352. doi: 10.1111/echo.14822

24. Gutor SS, Richmond BW, Du RH, Wu P, Lee JW, Ware LB, Shaver CM, Novitskiy SV, Johnson JE, Newman JH, et al. Characterization of Immunopathology and Small Airway Remodeling in Constrictive Bronchiolitis. Am J Respir Crit Care Med. 2022;206:260–270. doi: 10.1164/rccm.202109-2133OC

25. Galie N, Kim NH. Pulmonary microvascular disease in chronic thromboembolic pulmonary hypertension. Proc Am Thorac Soc. 2006;3:571–576. doi: 10.1513/pats.200605-113LR

26. Dorfmuller P, Gunther S, Ghigna MR, Thomas de Montpreville V, Boulate D, Paul JF, Jais X, Decante B, Simonneau G, Dartevelle P, et al. Microvascular disease in chronic thromboembolic pulmonary hypertension: a role for pulmonary veins and systemic vasculature. Eur Respir J. 2014;44:1275–1288. doi: 10.1183/09031936.00169113

27. Montani D, Lau EM, Dorfmuller P, Girerd B, Jais X, Savale L, Perros F, Nossent E, Garcia G, Parent F, et al. Pulmonary veno-occlusive disease. Eur Respir J. 2016;47:1518-1534. doi: 10.1183/13993003.00026-2016

28. Vachiery JL, Tedford RJ, Rosenkranz S, Palazzini M, Lang I, Guazzi M, Coghlan G, Chazova I, De Marco T. Pulmonary hypertension due to left heart disease. Eur Respir J. 2019;53. doi: 10.1183/13993003.01897-2018

29. Rosenkranz S, Howard LS, Gomberg-Maitland M, Hoeper MM. Systemic Consequences of Pulmonary Hypertension and Right-Sided Heart Failure. Circulation. 2020;141:678–693. doi: 10.1161/CIRCULATIONAHA.116.022362

30. Hassoun PM. Pulmonary Arterial Hypertension. N Engl J Med. 2021;385:2361-2376. doi: 10.1056/NEJMra2000348

31. Sandoval J, Salas J, Martinez-Guerra ML, Gomez A, Martinez C, Portales A, Palomar A, Villegas M, Barrios R. Pulmonary arterial hypertension and cor pulmonale associated with chronic domestic woodsmoke inhalation. Chest. 1993;103:12–20. doi: 10.1378/chest.103.1.12

32. Keusch S, Hildenbrand FF, Bollmann T, Halank M, Held M, Kaiser R, Kovacs G, Lange TJ, Seyfarth HJ, Speich R, et al. Tobacco smoke exposure in pulmonary arterial and thromboembolic pulmonary hypertension. Respiration. 2014;88:38–45. doi: 10.1159/000359972

33. Montani D, Lau EM, Descatha A, Jais X, Savale L, Andujar P, Bensefa-Colas L, Girerd B, Zendah I, Le Pavec J, et al. Occupational exposure to organic solvents: a risk factor for pulmonary veno-occlusive disease. Eur Respir J. 2015;46:1721–1731. doi: 10.1183/13993003.00814-2015

34. Sofianopoulou E, Kaptoge S, Graf S, Hadinnapola C, Treacy CM, Church C, Coghlan G, Gibbs JSR, Haimel M, Howard LS, et al. Traffic exposures, air pollution and outcomes in pulmonary arterial hypertension: a UK cohort study analysis. Eur Respir J. 2019;53. doi: 10.1183/13993003.01429-2018

35. Kaihara T, Yoneyama K, Nakai M, Higuma T, Sumita Y, Miyamoto Y, Watanabe M, Izumo M, Ishibashi Y, Tanabe Y, et al. Association of PM(2.5) exposure with hospitalization for cardiovascular disease in elderly individuals in Japan. Sci Rep. 2021;11:9897. doi: 10.1038/s41598-021-89290-5

36. Sangani R, Ghio A, Culp S, Patel Z, Sharma S. Combined Pulmonary Fibrosis Emphysema: Role of Cigarette Smoking and Pulmonary Hypertension in a Rural Cohort. Int J Chron Obstruct Pulmon Dis. 2021;16:1873–1885. doi: 10.2147/COPD.S307192

37. Chen WC, Park SH, Hoffman C, Philip C, Robinson L, West J, Grunig G. Right ventricular systolic pressure measurements in combination with harvest of lung and immune tissue samples in mice. J Vis Exp. 2013:e50023. doi: 10.3791/50023

38. Grunig G, Marsh LM, Esmaeil N, Jackson K, Gordon T, Reibman J, Kwapiszewska G, Park SH. Perspective: ambient air pollution: inflammatory response and effects on the lung’s vasculature. Pulm Circ. 2014;4:25–35. doi: 10.1086/674902

39. Liu J, Ye X, Ji D, Zhou X, Qiu C, Liu W, Yu L. Diesel exhaust inhalation exposure induces pulmonary arterial hypertension in mice. Environ Pollut. 2018;237:747–755. doi: 10.1016/j.envpol.2017.10.121

40. Haberzettl P, Conklin DJ, Abplanalp WT, Bhatnagar A, O’Toole TE. Inhalation of Fine Particulate Matter Impairs Endothelial Progenitor Cell Function Via Pulmonary Oxidative Stress. Arterioscler Thromb Vasc Biol. 2018;38:131–142. doi: 10.1161/ATVBAHA.117.309971

41. Dikalov S, Itani H, Richmond B, Vergeade A, Rahman SMJ, Boutaud O, Blackwell T, Massion PP, Harrison DG, Dikalova A. Tobacco smoking induces cardiovascular mitochondrial oxidative stress, promotes endothelial dysfunction, and enhances hypertension. Am J Physiol Heart Circ Physiol. 2019;316:H639–H646. doi: 10.1152/ajpheart.00595.2018

42. Gangwar RS, Bevan GH, Palanivel R, Das L, Rajagopalan S. Oxidative stress pathways of air pollution mediated toxicity: Recent insights. Redox Biol. 2020;34:101545. doi: 10.1016/j.redox.2020.101545

43. Riggs DW, Zafar N, Krishnasamy S, Yeager R, Rai SN, Bhatnagar A, O’Toole TE. Exposure to airborne fine particulate matter is associated with impaired endothelial function and biomarkers of oxidative stress and inflammation. Environ Res. 2020;180:108890. doi: 10.1016/j.envres.2019.108890

44. Almeida-Silva M, Cardoso J, Alemao C, Santos S, Monteiro A, Manteigas V, Marques-Ramos A. Impact of Particles on Pulmonary Endothelial Cells. Toxics. 2022;10. doi: 10.3390/toxics10060312

45. Hassoun PM, Mouthon L, Barbera JA, Eddahibi S, Flores SC, Grimminger F, Jones PL, Maitland ML, Michelakis ED, Morrell NW, et al. Inflammation, growth factors, and pulmonary vascular remodeling. J Am Coll Cardiol. 2009;54:S10–S19. doi: 10.1016/j.jacc.2009.04.006

46. Stacher E, Graham BB, Hunt JM, Gandjeva A, Groshong SD, McLaughlin VV, Jessup M, Grizzle WE, Aldred MA, Cool CD, et al. Modern age pathology of pulmonary arterial hypertension. Am J Respir Crit Care Med. 2012;186:261–272. doi: 10.1164/rccm.201201-0164OC

47. Hu Y, Chi L, Kuebler WM, Goldenberg NM. Perivascular Inflammation in Pulmonary Arterial Hypertension. Cells. 2020;9. doi: 10.3390/cells9112338

48. Mercurio V, Cuomo A, Naranjo M, Hassoun PM. Inflammatory Mechanisms in the Pathogenesis of Pulmonary Arterial Hypertension: Recent Advances. Compr Physiol. 2021;11:1805–1829. doi: 10.1002/cphy.c200025

49. Roos JE, McAdams HP, Kaushik SS, Driehuys B. Hyperpolarized Gas MR Imaging: Technique and Applications. Magn Reson Imaging Clin N Am. 2015;23:217–229. doi: 10.1016/j.mric.2015.01.003

