## Supplemental material for "Pulmonary Vascular Disease in Veterans with Post-Deployment Respiratory Syndrome"

**Supplemental Table 1.**

| Vessel compartment | ND controls | PDRS | PAH |
|---|---|---|---|
| BVB arteries | 297.6±32.4 | 299.7±38.4 | 304.7±32.6 |
| IA arteries | 98.6 (96.8, 112.1) | 105.1 (98.4, 114.9) | 120.2 (105.5, 127.4) |
| IA veins | 92.7 (85.2, 100.9) | 99.9 (91.4, 104.2) | 104.3 (90.9, 108.1) |

All differences between ND controls, PDRS and PAH patients were not significant.

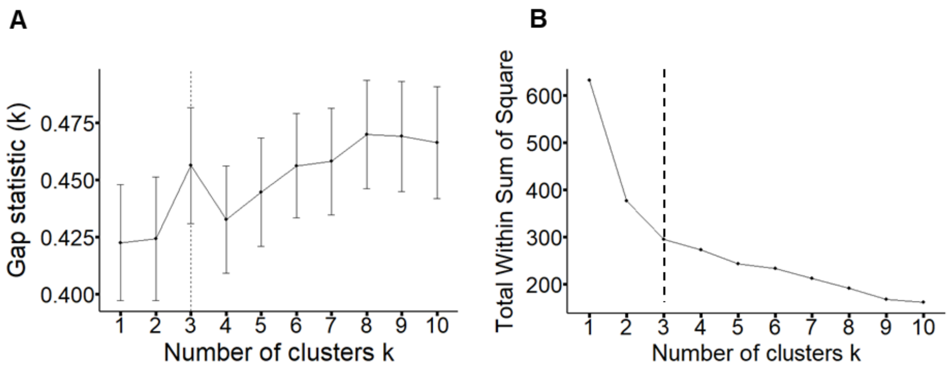


**Supplemental Figure 1.** A – Gap statistic (k) plot with a determination of an optimal number of clusters; B – Total Within Sum of Square plot with a determination of an optimal number of clusters. The optimal number of clusters is 3 by both used methods.

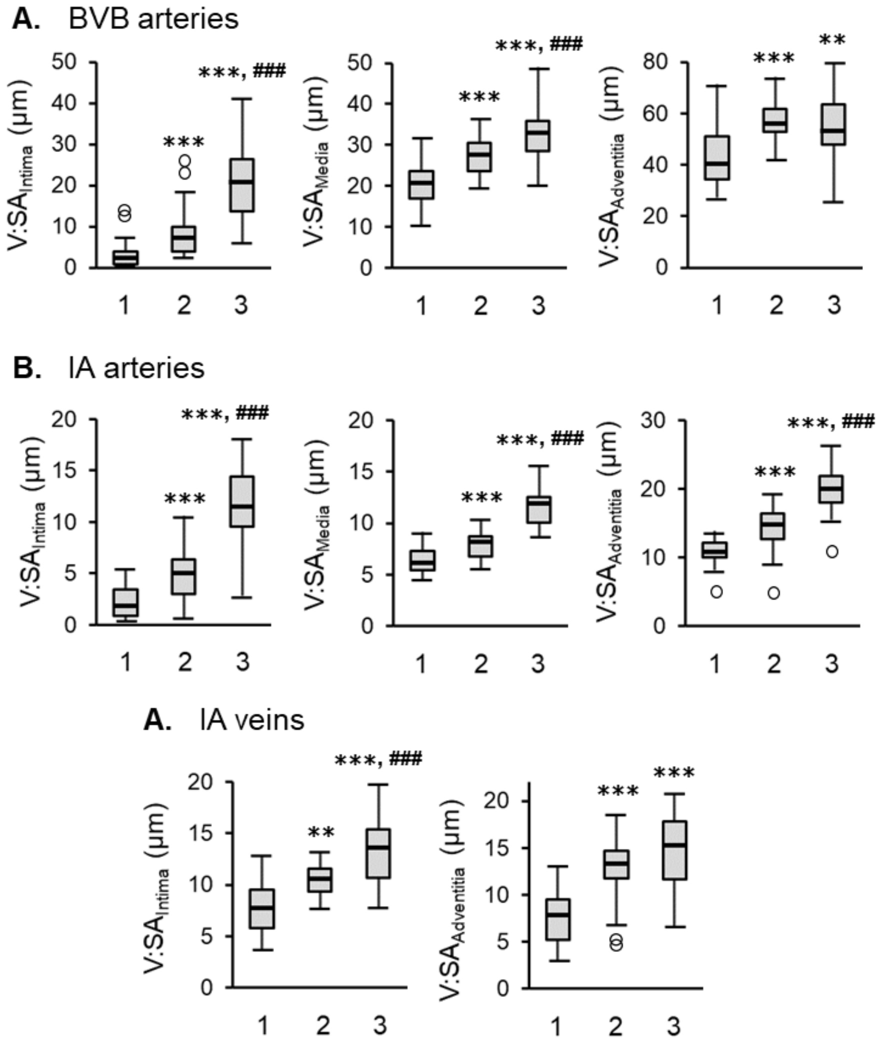


**Supplemental Figure 2.** *Comparison analysis of morphometrical data from soldiers with PDRS according to their clustering separation*. A-C – graph showing discrete increase of morphometrical parameters from 1$^{st}$ cluster towards 3$^{rd}$ cluster for BVB arteries (A), IA arteries (B) and IA veins (C). Data presented as V:SA$_{Intima}$ for intima thickness, V:SA$_{Media}$ for media thickness or V:SA$_{Adventitia}$ for adventitia thickness. Boxes represent the interquartile range, whiskers extend to the most extreme data point which is no more than 1.5 times the interquartile range from the box, and circles beyond the whiskers are extreme values, the line within the box represents the median. Groups were compared pairwise using Mann-Whitney U or Student t tests depending on variable distribution. Estimated p-values were Bonferroni-adjusted. *** – p<0.001 compared to ND controls; ## – p<0.01 compared to PDRS; ### – p<0.001 compared to PDRS.
